# INVESTIGATING TRAIT IMPULSIVITY IN OBSESSIVE-COMPULSIVE DISORDER: A SCOPING REVIEW

**DOI:** 10.1101/2024.10.02.24314767

**Authors:** M.J. Banwell, F. Scheffler, C. Lochner, S.R. Chamberlain, D.J. Stein

## Abstract

**Background/Aims:** While obsessive-compulsive disorder (OCD) is specifically characterised by compulsivity, considerable literature suggests impulsivity also plays an important role in the disorder. However, impulsivity is a multi-faceted construct and the exact relationship of trait impulsivity to OCD remains unclear. Therefore, this scoping review aimed to collate and review studies of trait impulsivity in OCD by an investigation of: 1. How trait impulsivity in OCD is measured in the research literature; 2. How people with OCD perform on these measures and compare to healthy and other psychiatric groups; and 3. What correlations with trait impulsivity are observed in OCD.

**Methods:** This study was pre-registered on PROSPERO (CRD42023481781). Electronic search of PubMed, Scopus, and PsycINFO databases using keywords ‘(impuls*) AND (OCD)’ was undertaken without date restrictions for peer-reviewed articles available in English. After exclusion of duplicates and screening of 1476 abstracts, 114 articles were identified for full-text review.

**Results:** 54 articles were reviewed after excluding studies assessing neurocognitive impulsivity only (i.e. no inclusion of trait impulsivity), sub-clinical OCD symptoms, and review articles. The literature reports cross-sectional clinician-rated and self-rated trait impulsivity data, with the Barratt Impulsivity Scale (BIS) being used most frequently. Broadly, people with OCD scored higher than healthy controls on at least one aspect of trait impulsivity. However, comparisons of OCD groups to other psychiatric groups demonstrated equal or lower trait impulsivity in OCD. Individuals with OCD with comorbid diagnoses (ADHD, behavioural addictions, tic disorder, borderline personality disorder, bipolar disorder) had relatively higher levels of trait impulsivity than those without. In OCD, trait impulsivity scores were associated with various psychiatric symptomatology (OCD severity, anxiety, depression, compulsivity, hoarding levels, behavioural addictions, anhedonia, aggressive and sexual impulses). Trait impulsivity did not correlate with neurocognitive measures of impulsivity.

**Conclusions:** Key findings are that trait impulsivity research in OCD was predominantly observational, with cross-sectional studies using the BIS. While higher levels of trait impulsivity were seen in patients with OCD compared to healthy controls, this finding was not specific to OCD. OCD demonstrated equal or lower trait impulsivity than other psychiatric groups that the literature had examined to date; trait impulsivity in OCD was positively correlated with a number of psychiatric factors; and neurocognitive measures of impulsivity did not correlate with trait impulsivity. Future work on OCD should include interventional and neuroimaging methods that utilise several different measures of impulsivity.

## Introduction

OCD is a prevalent neuropsychiatric disorder associated with substantial morbidity. In the first nationally representative survey using DSM-III criteria in the United States, OCD was the fourth most-common mental disorder^1^. Subsequent studies around the world have indicated an estimated lifetime prevalence of 2-3%^2,3^ and the Global Burden of Disease study has emphasised the high level of disability associated with OCD^4^. This is consistent with evidence that this chronic and relapsing condition exerts significant detrimental effects on individuals through loss of income, reduced quality of life, and heightened mortality risk^2,5–7^. On a societal level, these disruptions to individual functioning result in high economic costs, both through direct healthcare costs and indirect costs (e.g. productivity decline and sick leave)^8,9^.

Compulsivity and impulsivity are commonly co-occurring constructs that are represented in personality, cognition, and behaviour^10^. Compulsivity, a seminal characteristic of OCD^11^, can be defined as a habitual or stereotyped pattern of unwanted (overt or covert) behaviours enacted in line with a fixed set of rules or as attempts to avoid perceived negative outcomes^12^. Impulsivity is a broad construct that includes deficits in processes such as inhibitory control, self-regulation, decision-making, ‘craving’, situation-dependent impulsivity (i.e. state impulsivity), and trait impulsivity^13,14^. Trait impulsivity has been defined in various ways, including an individual’s relatively consistent propensity towards emotional or behavioural responses without deliberation or the evaluation of consequences^15^, which may take the form of ‘rapid, unplanned reactions to internal or external stimuli^16^,’ as well as insufficiently considered, risky, reckless, or prematurely enacted behaviours^17^. A recent meta-analysis^18^ addressed neurocognitive tasks of impulsivity in OCD, but there are few reviews of trait impulsivity in OCD as indexed using self-report questionnaires^19^.

In the absence of comprehensive collations of trait impulsivity in OCD, many questions remain – specifically, about the methods for assessing trait impulsivity in OCD, whether measures of impulsivity differ between individuals with OCD, individuals with other mental disorders, and healthy controls, and what factors are related to trait impulsivity in OCD. Thus, this review aimed to evaluate and synthesise: 1. How trait impulsivity in OCD is measured in the research literature; 2. How individuals with OCD perform on these measures in comparison to individuals with other mental disorders and healthy controls; and 3. What associations with trait impulsivity are observed in OCD.

## Methods

This study was pre-registered on PROSPERO (CRD42023481781). Search terms related to impulsivity and OCD (‘(impuls*) AND (OCD)’) were used to explore PubMed, Scopus, and PsycINFO databases. Search dates ranged from inception until the end of 2023. In order to identify any additional more recent literature that would have been missed by the original review, this process was repeated prior to submission on 20 September 2024. As shown in Figure 1, a total of 1 476 results were retrieved from the initial searches (PubMed – 444; Scopus – 451; PsycINFO – 581). After exclusion of duplicates and screening of abstracts, 114 articles were selected for full-text analysis, of which a total of 54 were included for review. Articles were analysed in terms of subject matter, aims, methods, results, and suggested conclusions. This data were extracted using a review matrix developed for the purpose of this review. Variables included: author(s)’s name(s), publication title, publication year, journal, location (data collection), sample size, recruitment strategy (e.g. mental health facility, university advertisement), type of data included (i.e. neurocognitive, neuroimaging, clinical), measure of trait impulsivity (name), conclusions, funders.

**Figure 1:**
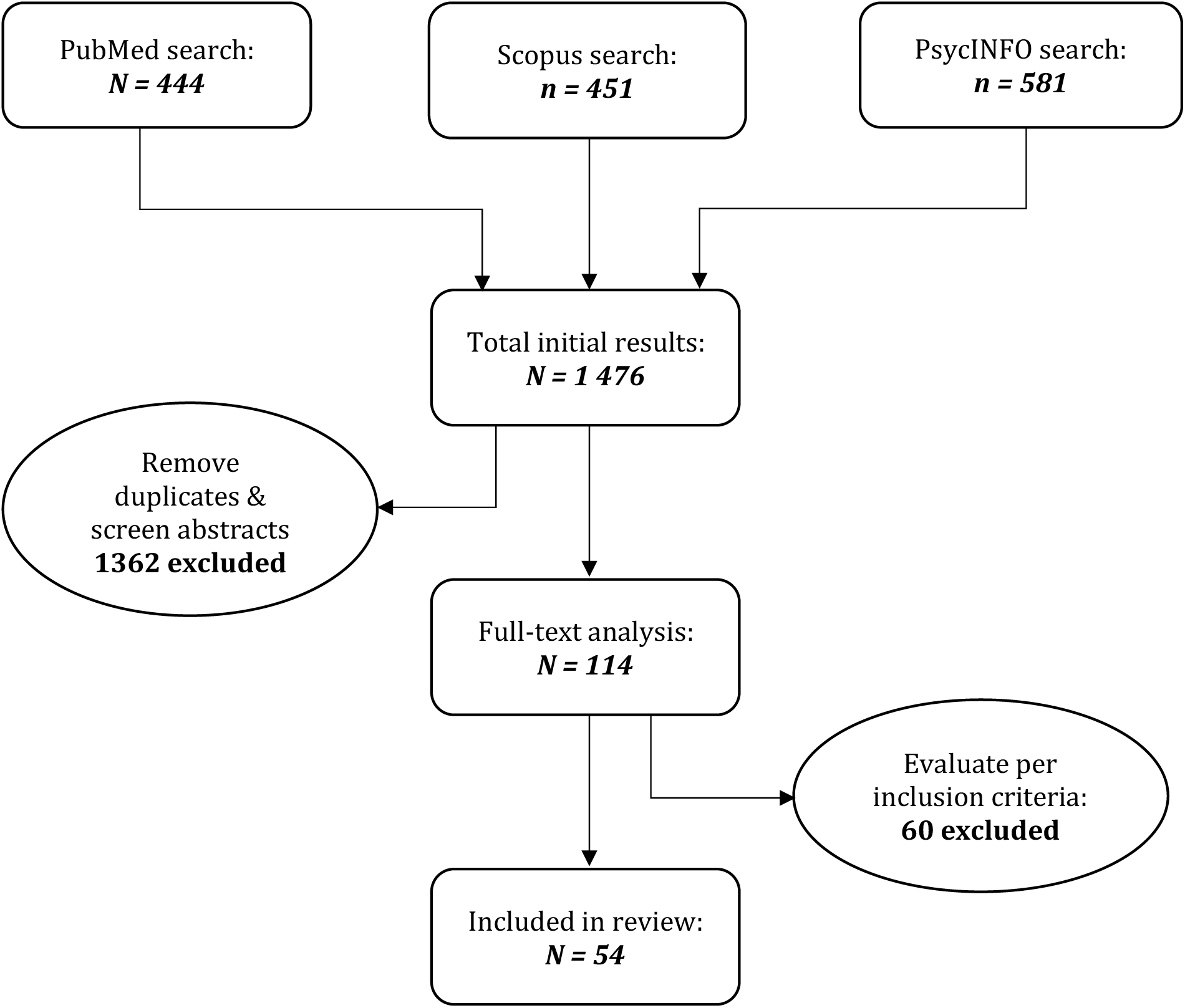
Flow diagram of review process.

### Inclusion/Exclusion Criteria

This review covered published, peer-reviewed studies, that are available in English, and that examined trait impulsivity in people with a primary diagnosis of OCD. This included research investigating only OCD participants, as well as those that used an OCD group as a comparative group. Given the broadness of the concept of impulsivity, it was decided to narrow the focus to trait impulsivity. As such, this review did not focus on more narrowly defined impulse control disorders (e.g. kleptomania) or more broadly defined impulse control disorders (e.g. substance use disorders), unless they were included as psychiatric control groups or comorbidities of OCD groups. Additionally, research assessing impulsivity as measured by neurocognitive tests (e.g. inhibitory control deficits) was only included when this was done in conjunction with measures of trait impulsivity.

## Results

As shown in Figure 1, a total of 1 476 results were retrieved from the initial searches (PubMed – 444; Scopus – 451; PsycINFO – 581). After exclusion of duplicates and screening of abstracts, 114 articles were selected for full-text analysis, of which a total of 54 were included for review. The most common reasons for exclusion were assessment of only neurocognitive impulsivity; review articles; full text not being available in English; and examination of obsessive-compulsive symptoms/features in community/psychiatric samples instead of participants diagnosed with OCD. A summary of the 54 reviewed articles and their findings and correlations is tabled in Appendix A. The secondary search conducted in September 2024 did not yield any new studies that met inclusion criteria for this review.

### Measuring trait impulsivity in OCD

Almost all studies (93%; *n* = 50) included male and female participants – four examined one sex (two all-male^20,21^, two all-female^22,23^). Predominantly, adults with OCD were studied (93%; *n* = 50), with youth (children and adolescents) being evaluated in four studies^24–27^, two of which used the same sample^24,26^. Exclusively outpatients were considered in 89% (*n* = 48) of studies – four included inpatients and outpatients, and two exclusively examined inpatients.

The Barratt Impulsivity Scale (BIS)^28^ was the primary measure for assessing trait impulsivity and was used in over 70% (*n* = 38) of studies. The Tridimensional Personality Questionnaire (TPQ)^29^ and a version of the UPPS Impulsive Behavior Scale^30^, respectively, were each used in three studies. Other measures, each used in one study, included the Plutchik Impulsivity Scale (PIS)^31^, Eysenck Psychoticism Scale^32^, Impulsive Behavior Scale (I8)^33^, Eysenck I-7^34^, and the Schalling Impulsivity Scale^35^. Impulsivity subscales were used in eight studies (Swanson, Nolan, and Pelham –IV Parent Scale [SNAP-IV]^36^, Childhood Suicide Potential Scale^37^, Scale for the Rapid Assessment of Psychopathology [SVARAD]^38^, PANDAS (paediatric autoimmune neuropsychiatric disorders associated with streptococcal infections) Questionnaire^39^, Karolinska Scales of Personality^40^, Eysenck Impulsiveness Venturesomeness Empathy Scale^41^, with two using the Eysenck Personality Questionnaire^42^). Finally, two studies developed their own measure of trait impulsivity – one^43^ using their own measure in conjunction with the Schalling Impulsivity Scale, and one^22^ using only their measure. Four studies included more than one measure of trait impulsivity^43–46^.

Samples wholly comprised of OCD participants were examined in 35% (*n* = 19) of studies. In just over half of these (*n* = 10), OCD participants were further split into comparative groups: smokers versus non-smokers, comorbidities, features of suicidal ideation, presence of childhood trauma, high versus low impulsivity or hoarding levels. Healthy controls (HC) were compared with an OCD group in 54% of the studies (*n* = 29), with 17 of these comparing only HC and OCD groups. Non-OCD psychiatric groups were included in 33.3% of studies (*n* = 18), with six exclusively evaluating psychiatric groups in comparison to an OCD group. Psychiatric groups included eating disorders (anorexia nervosa, bulimia nervosa), anxiety disorders (panic disorder, social phobia), affective disorders (major depressive disorder, bipolar disorder) impulse control disorders (pathological gambling, compulsive buying), OCD-related disorders (skin-picking, trichotillomania, tic disorders), borderline personality disorder, schizophrenia, conduct disorder, and attention deficit hyperactivity disorder (ADHD).

### Reported Findings

#### OCD vs. HC

Higher scores were observed in OCD groups than HC on at least one dimension of trait impulsivity in 83% of studies that included HC (*n* = 25). Of the studies indicating elevated trait impulsivity in OCD compared to HC, almost 70% (*n* = 17) reported this difference in total BIS scores; 60% (*n =* 15) in BIS attention scores; and 24% (*n* = 6) in BIS non-planning subscale. Non-planning impulsivity was only significantly elevated in OCD compared to HC when BIS total and/or attention were also significantly greater. A single study^47^ indicated an elevated motor subscale – people with OCD who exhibited high levels of hoarding demonstrated a higher motor subscale than HC, who were equal to OCD participants with low levels of hoarding. Three studies^20,48,49^ reported no differences between healthy control groups and OCD groups in all aspects of trait impulsivity, while two studies^24,26^ reported lower trait impulsivity levels in OCD participants than HC.

#### OCD vs Other Psychiatric Groups

Over 60% (*n* = 11) of the 18 studies comparing OCD and psychiatric controls reported no difference between OCD groups and other psychiatric groups on various measures of trait impulsivity. These samples included patients with eating disorders (indicated by BIS total, PIS, and a measure developed for study^22^), psychotic disorders (PIS; UPPS total and premeditation, perseverance, and sensation-seeking subscales), pathological gambling (PG) (BIS total [two studies]; BIS non-planning and motor subscales), affective disorders (BIS total, PIS), social phobia and panic disorder (BIS total), skin-picking and trichotillomania (Schalling Impulsivity Scale), conduct disorders (PIS), and alcohol dependence (BIS total).

Almost 40% (*n* = 7) of these 18 studies reported OCD groups displaying significantly lower trait impulsivity scores than other psychiatric groups on several measures. These included groups of participants with ADHD (IVE impulsivity and risk-taking), compulsive buying (BIS total and all subscales), trichotillomania (Eysenck I-7 impulsiveness subscale), PG (BIS non-planning, TPQ, an impulsivity phenotype derived from the UPPS-P, and an impulsivity composite derived from the TRQ and correlated with BIS), and borderline personality disorder (UPPS total, urgency, premeditation; Eysenck I-7, TPQ). While participants with schizophrenia^50^ scored significantly lower than OCD participants on the UPPS Urgency subscale, UPPS total and other subscales were lower in the OCD group than the schizophrenia group.

#### OCD vs. OCD

A total of 23 studies split an OCD population into sub-groups to compare trait impulsivity. Three examined smoking status^51–53^, four assessed comorbidity of ADHD^54–57^, seven investigated other comorbidities that were not depression and/or anxiety (PANDAS^25^, behavioural addictions^58^, eating disorders^22^, personality disorders^59^, bipolar disorder^60^, tic disorder^61^, hoarding features^47^), three evaluated features of suicidality^62–64^, and three analysed symptom profiles of OCD^65–67^. The remaining three examined childhood trauma (CT)^68^, sex differences^69^, and high-impulsivity versus low-impulsivity^44^, respectively.

Across studies, OCD groups with respective comorbidities displayed elevated trait impulsivity scores compared to those without them (ADHD, PANDAS, behavioural addictions, borderline personality disorder, bipolar disorder, tic disorder, high levels of hoarding), except in the case of obsessive-compulsive personality disorder (OCPD). OCD with OCPD participants showed significantly reduced BIS total and non-planning scores than those with only OCD. Trait impulsivity in people with OCD was also observed to be higher in OCD groups that were smokers compared to non-or former smokers, and in those with comorbid bulimia compared to those with comorbid anorexia. Mixed results were reported when comparing OCD participants who had a lifetime history of suicide attempts (SA), suicidal ideation (SI), or no suicidal history (NS). One study^62^ found no differences in trait impulsivity between these groups, another^64^ found that participants with SA scored higher on BIS attention but lower on BIS non-planning than the other groups, while the third^63^ found that OCD participants with comorbid depression with lifetime history of SI scored higher on trait impulsivity than those without SI history.

OCD groups with sexual and aggressive impulses demonstrated higher trait impulsivity than those with other primary symptoms^65,66^, but trait impulsivity did not differ across washing, taboo thoughts, and symmetry/counting/repeating/ordering dimensions^67^. Regarding sex differences, only BIS attention was elevated in males with OCD as compared to females (OCD and controls).

### Correlations with trait impulsivity in OCD

Correlational results of all studies are tabled in Appendix A. Broadly speaking, trait impulsivity significantly positively correlated with obsessive-compulsive symptom presence and severity; depression; anxiety; behavioural addictions (internet/sex/pornography/ mobile phone/kleptomania/pyromania/gambling/food/exercise) and childhood trauma. Trait impulsivity was negatively correlated with conscientiousness. Mixed results were observed for the correlation of trait impulsivity with hoarding symptoms. Trait impulsivity was not found to correlate significantly with treatment outcome, nor with suicidal behaviour in OCD adolescent inpatients. Furthermore, every study that examined correlations between trait impulsivity and neurocognitive impulsivity measures in OCD reported non-significant results^19,20,23,69–71^.

## Discussion

This scoping review considered investigations measuring trait impulsivity in OCD and their comparisons with healthy and other psychiatric groups. The main findings were: 1. Trait impulsivity in OCD was primarily assessed using the BIS; 2. OCD groups largely demonstrated elevated levels of trait impulsivity in comparison to HC groups; 3. OCD groups exhibited similar or decreased trait impulsivity in comparison to most psychiatric groups; 4. Several psychiatric symptoms significantly positively correlated with trait impulsivity; and 5. Trait impulsivity scores did not correlate significantly with any measures of neurocognitive impulsivity.

Almost all of the 54 reviewed studies conducted cross-sectional observational research in adult male and female outpatients with OCD. There were few interventional and neuroimaging studies examining trait impulsivity in OCD. The vast majority of these studies (exceeding 70%) used the BIS to measure trait impulsivity in OCD. While the BIS is considered the gold standard of trait impulsivity measurement^72^, questions regarding the psychometric properties of the current version (BIS-11), including factor structure, have emerged (see, for example, Reise et al., 2013^72^; Kapitány-Fövény et al., 2020^73^; Kahn et al. 2019,^74^). Future work may consider including the BIS alongside alternative measures of trait impulsivity (such as the short-form version, BIS-15^75^) and in conjunction with measures of other forms of impulsivity (such as the SUPPS-P^76^, which has been found to retain good psychometric properties and assess various domains of impulsivity while being less time-consuming than the original format^77^) in order to ascertain a clearer clinical picture of impulsivity.

Most research found an elevated score on trait impulsivity in OCD groups compared to HC groups. It is notable that in one study that did not find this, all OCD participants were receiving psychiatric mediation. While psychiatric medication has been associated with decreased self-report trait impulsivity in other psychiatric conditions^78,79^, other research in this review found higher trait impulsivity in OCD participants on medication than in HC^19,69,70,80^. In contrast to most OCD groups exhibiting higher levels of trait impulsivity than HC, the literature was consistent in reporting that individuals with other mental disorders have equal or higher levels of trait impulsivity than individuals with OCD. Taken together, these results underscore that elevated trait impulsivity in comparison to HC is not specific to OCD, but is found across a range of psychiatric conditions.

Trait impulsivity was higher in OCD groups that had other psychiatric comorbidities (excluding OCPD), those that also had bulimia in comparison to anorexia, who were smokers, and those who had primary symptoms of sexual and aggressive thoughts/impulses towards themselves and others. Mixed results were observed when comparing OCD groups with varying levels of current and lifetime suicidal ideation and attempts. Sex differences were only found in BIS attention, with males with OCD demonstrating elevated scores compared to all controls and females with OCD. These results suggest that while OCD was associated with higher levels of trait impulsivity than HC, this effect was augmented when psychiatric comorbidities are present.

Positive correlational results in this review provided further evidence for the co-occurrence of psychiatric symptoms and trait impulsivity in OCD. In OCD participants, positive correlations with trait impulsivity were observed in those with comorbid psychiatric disorders. These include depression, anxiety, and behavioural addictions. Furthermore, positive correlations also emerged when considering obsessive-compulsive symptoms of aggressive thoughts/impulses towards the self/others, cleaning, checking, contamination, and symmetry. Symptom severity also yielded a positive correlation with trait impulsivity in OCD. Given these associations, future psychiatric interventions may consider targeting trait impulsivity in treatment. These results contrast with a recent meta-analysis^18^ which found impulsivity did not vary according to a number of clinical factors (medication status, presence of comorbidities, OCD symptom severity, illness duration). However, the meta-analysis only investigated neurocognitive measures of impulsivity, which did not correlate with trait impulsivity in our review.

Notably, every study that examined correlations between trait impulsivity and neurocognitive impulsivity measures in OCD reported non-significant results^19,20,23,69–71^. One study reported the same absence of correlations with trait impulsivity in HC^70^. These findings are consistent with previous work emphasising that self-report surveys and behavioural tasks are operating at different levels of measurement, and even when surveys and tasks pertain to the same psychological construct, they may not correlate^81^. They underscore the heterogeneity of impulsivity as a construct, and the need to measure impulsivity using multidimensional assessment^71^. It is conceivable, also, that poor inhibitory control observed in OCD on neurocognitive tests of impulsivity may reflect severity of symptoms rather than an inherent response suppression deficit^20^. People with OCD may consider themselves as more impulsive in general, particularly when their symptoms are more severe^20^ and correlations between OCD symptom severity and trait impulsivity may offer a tentative explanation for the dissociation between trait and neurocognitive impulsivity measures.

Limitations of the study included being unable to access English versions of some studies. The design of this study was to focus on trait impulsivity, and this means that other key aspects of impulsivity, such as impulse control disorders, or neurocognitive tests of impulsivity, were not addressed in detail.

In summary, a number of conclusions can be drawn about trait impulsivity in OCD. Trait impulsivity research in OCD has been predominantly observational, with cross-sectional studies using the BIS most frequently. People with OCD tended to have higher trait impulsivity scores than healthy controls, but equal or lower scores than individuals with several other mental disorders, suggesting that trait impulsivity is not unique to OCD. In people with OCD, positive correlations were observed between trait impulsivity and OCD severity, anxiety, depression, and behavioural addictions, suggesting that interventions targeting trait impulsivity may improve treatment outcomes in OCD. Trait impulsivity scores did not, however, correlate with measures of neurocognitive impulsivity. Future work on OCD should include interventional and neuroimaging research, and the effect of medication on trait impulsivity should be further explored.

## Data Availability

All data produced in the present study is included in Appendix A.

## Financial Support

This review was made possible (in part) by a grant from Carnegie Corporation of New

York (MJB, grant number G-21-58838). The statements made and views expressed are solely the responsibility of the author.

CL and DJS are supported by the SAMRC.

SRC and CL receive a stipend from Elsevier for journal editorial work.

DJS has received consultancy honoraria from Discovery Vitality, Johnson & Johnson, Kanna, L’Oreal, Lundbeck, Orion, Sanofi, Servier, Takeda and Vistagen.

All authors declare that they have no conflict of interest.

## Appendix A: Studies Included in this Review

| Reference | Location | Sample Size | Recruitment Strategy | Type of Data | Type of Study | Impulsivity Measure | Conclusions | Limitations | Correlations |
| --- | --- | --- | --- | --- | --- | --- | --- | --- | --- |
| Abramovitch, A., Dar, R., Hermesh, H. & Schweiger, A. Comparative neuropsychology of adult obsessive-compulsive disorder and attention deficit/hyperactivity disorder: Implications for a novel executive overload model of OCD. *J. Neuropsychol.* **6**, 161–191 (2012). | Israel | 30 OCD<br>30 ADHD<br>30 HC | MH facility OCD; college LD support centre ADHD; RA network for HC | Clinical/<br>NC | Obs | Eysenck IVE impulsivity and venturesomeness subscales | ADHD > OCD IVE impulsivity and risk-taking. OCD – HC IVE impulsivity. HC > OCD IVE risk-taking. | +/- 50% OCD sample medicated; ADHD group unmedicated. Only male participants. | IVE impulsiveness significantly positively correlated with OCI-R scale of OCD symptom severity only in OCD group. |
| Apter, A. *et al.* Depression and suicidal behavior in adolescent inpatients with obsessive compulsive disorder. *J. Affect. Disord.* **75**, 181–189 (2003). | Israel | 40 OCD<br>118 PsyD<br>53 AD<br>81 CD<br>50 ED<br>87 HC | MH facility; HC marketing service | Clinical | Obs | Plutchik Impulsivity Scale | HC > OCD impulsiveness. OCD = patient population impulsivity. Discussion stated OCD inpatients have high levels of impulsivity – unclear from results where this conclusion emerged. | Only inpatients. | Impulsiveness significantly positively correlated with levels of suicidal behaviour in patient controls, but not OCD. |
| Bejerot, S., von Knorring, L. & Ekselius, L. Personality traits and smoking in patients with obsessive-compulsive disorder. *Eur. Psychiatry J. Assoc. Eur. Psychiatr.* **15**, 395–401 (2000). | Sweden | 64 OCD<br>(51 non-smokers, 13 smokers) | MH facility; drug trial; OCD association | Clinical | Obs | Karolinska Scales of Personality – Impulsiveness subscale | Smoking OCD > non-smoking OCD impulsivity. | Size discrepancy between groups. |  |
| Benatti, B. *et al.* Prevalence and Correlates of Current Suicide Risk in an International Sample of OCD Adults: A Report from the International College of Obsessive-Compulsive Spectrum Disorders (ICOCS) Network and Obsessive | Not stated (presume Italy) | 75 OCD<br>70 HC | MH facility OCD; HC not stated | Clinical | Obs | BIS | OCD > HC BIS total and attentional impulsivity; true for pure and comorbid patients. |  |  |
| Compulsive and Related Disorders Network (OCRN) of the European College of Neuropsychopharmacology. *J. Psychiatr. Res.* **140**, 357–363 (2021). |  |  |  |  |  |  |  |  |  |
| Bernstein, G. A., Victor, A. M., Pipal, A. J. & Williams, K. A. Comparison of clinical characteristics of pediatric autoimmune neuropsychiatric disorders associated with streptococcal infections and childhood obsessive-compulsive disorder. *J. Child Adolesc. Psychopharmacol.* **20**, 333–340 (2010). | USA | 39 OCD (21 PANDAS, 18 non-PANDAS) | MH facility | Clinical | Obs | PANDAS Questionnaire items: inattention/ hyperactivity/ impulsivity | Children with PANDAS OCD are more likely to demonstrate impulsivity in their initial episode of neuropsychiatric illness based on parent report. | Parents retroactively reported symptoms. Limited measure. |  |
| Boisseau, C. L. *et al.* Behavioral and cognitive impulsivity in obsessive-compulsive disorder and eating disorders. *Psychiatry Res.* **200**, 1062–1066 (2012). | USA | 19 OCD<br>21 ED<br>21 HC | MH facility | Clinical/ NC | Obs | BIS | OCD = ED > HC BIS impulsivity. Only OCD demonstrated elevated stop-signal reaction time. | Only female sample. |  |
| Bottesi, G., Ghisi, M., Ouimet, A. J., Tira, M. D. & Sanavio, E. Compulsivity and Impulsivity in Pathological Gambling: Does a Dimensional-Transdiagnostic Approach Add Clinical Utility to DSM-5 Classification? *J. Gambl. Stud.* **31**, 825–847 (2015). | Italy | 44 PG<br>75 AUD<br>22 OCD<br>75 HC | MH facility; HC community | Clinical/ NC | Obs | BIS | OCD = PG and AUD > HC on BIS total. OCD < all other groups on NC tests. | Only BIS total used. |  |
| Çoban, A. & Tan, O. Attention Deficit Hyperactivity Disorder, Impulsivity, Anxiety, and Depression Symptoms Mediating the Relationship Between Childhood Trauma | Turkey | 106 OCD (60 CT<br>46 no CT) | MH facility | Clinical | Obs | BIS | OCD CT > OCD no CT BIS total and subscales. Seems trauma increases impulsivity levels in patients who are already impulsive. |  | BIS non-planning significantly weakly negatively correlated with |
| and Symptoms Severity of Obsessive-Compulsive Disorder. *Noro Psikiyatri Arsivi* **57**, 37–43 (2020). |  |  |  |  |  |  |  |  | Y-BOCS Compulsion. |
| Demartini, B. *et al.* Reduced interoceptive accuracy in patients with obsessive-compulsive disorder: A case-control study. *J. Clin. Neurosci. Off. J. Neurosurg. Soc. Australas.* **90**, 152–154 (2021). | Italy | 18 OCD<br>18 HC | MH facility | Clinical | Obs | BIS | OCD = HC | Small sample size. |  |
| Rai, D., Jaisoorya, T. S., Narayanaswamy, J. C., Arumugham, S. S., & Reddy, Y. J. (2022). Behavioural addictions in obsessive compulsive disorder—Prevalence and clinical correlates. *Psychiatry Research Communications*, *2*(1), 100016. | India | 150 OCD<br>131 HC | MH facility; HC community sample | Clinical | Obs | BIS | OCD > HC BIS total. OCD with BA > other OCD BIS total, motor, and attention. | As criteria of most BA undefined, diagnosis was not established and 'risk for BA' may be more appropriate here. | BIS positively correlated with all BAs except CB in whole sample. |
| Ettelt, S., Ruhrmann, S., Barnow, S., Buthz, F., Hochrein, A., Meyer, K., ... & Grabe, H. J. (2007). Impulsiveness in obsessive-compulsive disorder: results from a family study. *Acta Psychiatrica Scandinavica*, *115*(1), 41-47. | Germany | 70 OCD + 139 relatives, 70 matched HC + 134 relatives | MH facility; cross-sectional population study | Clinical | Obs | BIS | OCD > HC BIS attention. Relatives OCD = relatives HC on BIS. | Medication status not stated. | Cognitive impulsivity positively correlated with symptoms of aggressive thoughts and impulses, checking, and symmetry. This was augmented when only considering OCD without comorbid diagnoses, as well as with contamination symptoms and PADUA total. In |
|  |  |  |  |  |  |  |  |  | OCD and relatives, age was significantly related to BIS total and non-planning. In relatives, sex had a significant effect on BIS total. |
| Ferrão, Y. A., Almeida, V. P., Bedin, N. R., Rosa, R., & Busnello, E. D. A. (2006). Impulsivity and compulsivity in patients with trichotillomania or skin picking compared with patients with obsessive-compulsive disorder. *Comprehensive psychiatry*, 47(4), 282-288. | Brazil | 20 OCD<br>20 SPD/TTM | MH facility | Clinical | Obs | Schalling Impulsivity Scale; Multidimensional Impulsive-Compulsive Spectrum Assessment Instrument designed for this study | OCD = SPD/TTM on SIS. On developed measure: OCD believed they could delay impulses while SP/TTM believe they occur suddenly. OCD will spend time thinking about will act while SP/TTM take action as soon as possible.<br>OCD felt guilt over acting out while SP/TTM felt indifferent/ pleased.<br>At the time of acting out, OCD felt guilt where SP/TTM felt indifferent or pleased. After the act, both groups felt guilt.<br>OCD has rituals when acting out, unlike SP/TTM. OCD believed harm may come if they are<br>prevented from acting, SP/TTM believe they may benefit if not able to act. | Nonmatched groups. Half OCD group had comorbidities (MDD 7, AUD 2, TS 1). |  |
| Filomensky, T. Z., Almeida, K. M., Nogueira, M. C. C., Diniz, J. B., Lafer, B., Borcato, S., & Tavares, H. (2012). Neither bipolar nor obsessive-compulsive disorder: compulsive buyers are impulsive acquirers. *Comprehensive Psychiatry*, 53(5), 554-561. | Brazil | 35 OCD<br>24 CB<br>21 BD | MH facility | Clinical | Obs | BIS | CB > OCD on BIS total and all subscales. Differences not reported between OCD and BD-1. |  |  |
| Frydman, I., Mattos, P., de Oliveira-Souza, R., Yücel, M., Chamberlain, S. R., Moll, J., & Fontenelle, L. F. (2020). Self-reported and neurocognitive impulsivity in obsessive-compulsive disorder. *Comprehensive Psychiatry*, 97, 152155. | Brazil | 17 OCD<br>17 HC | MH facility;<br>institute support staff | Clinical | Obs | BIS | OCD > HC BIS total and attention despite no differences in neurocognitive impulsivity. OCD may see selves as more impulsive (self-report), which correlates to severity of OCD symptoms. Note: all OCD medicated. | Very small sample - could only detect large group effect sizes. Corrections for multiple comparisons not performed. Didn't rule out history of psychiatric disorders in HC. | OCI-R scores predicted BIS attention scores. |
| Fullana, M. A., Mataix-Cols, D., Caseras, X., Alonso, P., Menchón, J. M., Vallejo, J., & Torrubia, R. (2004). High sensitivity to punishment and low impulsivity in obsessive-compulsive patients with hoarding symptoms. *Psychiatry research*, 129(1), 21-27. | Spain | 56 OCD with hoarding symptoms. | MH facility | Clinical | Obs | Eysenck's Psychoticism Scale | Hoarding symptoms associated with higher scores on Gray's Sensitivity to Punishment Scale and lower scores on EPQ. | No control group of OCD without hoarding symptoms. Used Y-BOCS checklist as hoarding indicator, which is not designed to be used as a scale. | Negative correlation between hoarding symptoms in OCD and Eysenck's Psychoticism scale. |
| Grassi, G., Makris, N., & Pallanti, S. (2020). Addicted to compulsion: assessing three core dimensions of addiction across obsessive-compulsive disorder and gambling disorder. *CNS spectrums*, 25(3), 392-401. | Italy | 44 OCD<br>26 PG<br>40 HC | MH facility, advertisements, word of mouth | Clinical/NC | Obs | BIS | OCD = GD > HC BIS total and BIS attention.<br>OCD = GD = HC BIS motor and BIS non-planning. | Authors noted that they did not assess functioning and disability, which may impact group scores. Groups differed on level of education and IQ (PG < OCD and HC). | In OCD group, BIS total positively correlated with HAM-D, HAM-A, and SHAPS anhedonia score; not true for PG. No mention of BIS correlating with IGT results. |
| Grassi, G., Pallanti, S., Righi, L., Figuee, M., Mantione, M., Denys, D., ... & Stratta, P. (2015). Think twice: Impulsivity and decision making in obsessive-compulsive disorder. *Journal of behavioral addictions*, 4(4), 263-272. | Italy | 38 OCD<br>39 HC | MH facility | Clinical/NC | Obs | BIS | OCD > HC BIS total, BIS attentional, and BIS non-planning. | Exclusion of comorbidities limits generalisability to general OCD population. | No significant correlations between clinical variables. Regression analysis showed medication didn't influence results. |
| Gupta, G., Avasthi, A., Grover, S., & Singh, S. M. (2014). Factors associated with suicidal ideations and suicidal attempts in patients with obsessive compulsive disorder. *Asian journal of psychiatry*, 12, 140-146. | India | 130 OCD<br>(30 lifetime SI with MDD<br>51 lifetime SI without MDD<br>49 without SI) | MH facility | Clinical | Obs | BIS | Current depression and current SI: no difference between groups (OCD with depression and SI, OCD without depression with SI, OCD without depression without SI).<br>Lifetime SI and current depression: OCD with lifetime SI<br>with and without depression > OCD without lifetime SI.<br>Lifetime SI and lifetime depression: OCD with lifetime SI and lifetime depression > OCD without lifetime SI.<br>Concluded Impulsivity may be risk factor for lifetime, but not current, SI. |  |  |
| Hoehn-Saric, R., & Barksdale, V. C. (1983). Impulsiveness in obsessive-compulsive patients. *The British journal of psychiatry*, 143(2), 177-182. | USA | 20 OCD (10 impulsive, 10 non-impulsive) | MH facility | Clinical | Obs | BIS, Eysenck Personality Questionnaire Form A | Impulsive OCD > non-impulsive group: neuroticism, HADS-D, BIS (total and all sub-scales), pathological childhood disturbances. | OCD diagnostic measure not reported. Impulsive measure is very dated – refers to vandalism, sexual assault, violence, substance use, homosexual acts. |  |
| Hwang, J. Y., Shin, Y. C., Lim, S. W., Park, H. Y., Shin, N. Y., Jang, J. H., ... & Kwon, J. S. (2012). Multidimensional comparison of personality characteristics of the big five model, impulsiveness, and affect in pathological gambling and obsessive-compulsive disorder. *Journal of* | Korea | 18 OCD<br>15 PG<br>33 HC | MH facility, HC internet advertisement | Clinical | Obs | BIS | BIS total: PG = OCD. OCD vs HC not significant, but trending to higher. OCD = PG > HC BIS attention. PG > OCD BIS non-planning. | Only male sample. Didn't control for mood disorders as a covariate. Generalisability may be limited -other psychiatric disorders were excluded | Conscientiousness negatively correlated with all factors of BIS; BDI positively correlated with non-planning impulsiveness; BAI positively correlated with |
| *Gambling Studies*, 28, 351-362. |  |  |  |  |  |  |  | (substance use, neurological, psychotic, TBI). | motor impulsivity. |
| Ibrahim, L., Abouhendy, W., Raafat, N., & Fouad, A. A. (2020). Prevalence and correlates of attention deficit hyperactivity disorder in obsessive-compulsive disorder patients. *Middle East Current Psychiatry*, 27, 1-8. | Egypt | 100 OCD | MH facility | Clinical | Obs | BIS | OCD with and without adult ADHD: OCD with adult ADHD > OCD without adult ADHD BIS total, non-planning, motor.<br>OCD with childhood ADHD, with and without adult ADHD: No difference BIS attention, OCD severity, depression, or anxiety.<br>OCD with childhood but not adult ADHD > OCD without lifetime ADHD BIS attentional. | Small sample size. Relies on retroactive recall of childhood symptoms. |  |
| Kim, S. T., Seo, J. H., Park, C. I., Kim, H. W., Boo, Y. J., Kim, H., ... & Kim, S. J. (2023). Core clinical symptoms and suicidal ideation in patients with obsessive-compulsive disorder: A network analysis. *Psychiatry and Clinical Neurosciences*, 77(2), 110-117. | Korea | 444 OCD<br>228 HC | MH facility, community advertisement | Clinical | Obs | BIS | OCD > HC BIS total. While impulsivity was not directly associated with suicidal ideation; it showed connections with alexithymia, unacceptable thoughts, and lassitude, which were directly related to suicidal ideation. | Did not report BIS subscale results. |  |
| Koenn, L. G., Kohl, S., Schleyken, S., & Kuhn, J. (2023). Impulsivity and | Germany | 16 OCD<br>14 TS<br>28 HC | MH facility, HC were subjects in previous studies | Clinical/NC | Obs | BIS | OCD > HC only BIS attention. OCD = TS. Behavioural testing | Small N, heterogeneity in severity of |  |
| Attention in Obsessive Compulsive and Tic Disorders: Mismatch in Self-Report and Behavioural Data. *Journal of Clinical Medicine*, 12(6), 2277. |  |  |  |  |  |  | (IMT/DMT) showed no difference between groups. | disease, more male than female, sex of HC not reported. |  |
| Martin, S., Graziani, P., & Del-Monte, J. (2021). Comparing impulsivity in borderline personality, schizophrenia and obsessional-compulsive disorders: Who is ahead?. *Journal of Clinical Psychology*, 77(7), 1732-1744. | France | 25 OCD<br>23 SZD<br>50 BPD<br>24 HC | MH facility, HC associative networks | Clinical | Obs | UPPS | BPD > OCD UPPS total, urgency, lack of premeditation. OCD = HC and schizophrenia UPPS total and lack of premeditation. OCD > HC and schizophrenia UPPS urgency. OCD = all groups lack of perseverance and sensation-seeking. | Non-homogenous groups in terms of sex (no males BPD), OCD higher depression rates. Small sample. | No correlation found in OCD patients for UPPS and clinical scores. |
| Martoni, R. M., De Filippis, R., Cammino, S., Giuliani, M., Risso, G., Cavallini, M. C., & Bellodi, L. (2018). Planning functioning and impulsiveness in obsessive-compulsive disorder. *European archives of psychiatry and clinical neuroscience*, 268, 471-481. | Italy | 68 OCD<br>68 HC | MH facility | Clinical/NC | Obs | BIS | OCD > HC BIS total and non-planning. OCD males > females (OCD and controls) BIS attention. |  | No correlation impulsivity scores and planning abilities. |
| Matsunaga, H., Miyata, A., Iwasaki, Y., Matsui, T., Fujimoto, K., & Kiriike, N. (1999). A comparison of clinical features among Japanese eating-disordered women with obsessive-compulsive disorder. *Comprehensive Psychiatry*, 40(5), 337-342. | Japan | 18 OCD<br>'pure'<br>16 OCD/AN<br>16 OCD/BN<br>16 OCD/AN & BN | MH facility | Clinical | Obs | Own measure previously used in another study. | OCD = ED group. OCD with BN > OCD with AN. | Developed own measure (i.e. not validated). |  |
| Melca, I. A., Yucel, M., Mendlowicz, M. V., de Oliveira-Souza, R., & | Brazil | 110 OCD | MH facility | Clinical | Obs | BIS | OCD without OCPD > OCD with OCPD BIS | Multiple comparisons - |  |
| Fontenelle, L. F. (2015). The correlates of obsessive–compulsive, schizotypal, and borderline personality disorders in obsessive–compulsive disorder. *Journal of anxiety disorders*, 33, 15-24. |  |  |  |  |  |  | non-planning and total.<br>OCD with SPD = OCD without SPD all BIS measures.<br>OCD with BPD > OCD without BPD<br>BIS total, motor, non-planning. | inflated type 1 error possible. |  |
| Memis, C. O., Dogan, B., Sevincok, D., Tunagur, T., Memis, S. D., & Sevincok, L. (2020). Which factors may differentiate lifetime suicide attempters from ideators in obsessive–compulsive disorder patients?. *Indian journal of psychiatry*, 62(4), 392-399. | Turkey | 160 OCD | MH facility | Clinical | Obs | BIS | No difference between groups who had lifetime SI, lifetime SA, and non-suicidal. | Relied on retrospective recall. Didn't compare comorbidities in OCD. Methods and frequency of SA not explored - only looked at one or more. |  |
| Mersin Kilic, S., Dondu, A., Memis, C. O., Ozdemiroglu, F., & Sevincok, L. (2020). The clinical characteristics of ADHD and obsessive-compulsive disorder comorbidity. *Journal of attention disorders*, 24(12), 1757-1763. | Turkey | 67 OCD (32 with ADHD, 35 without ADHD) | MH facility | Clinical | Obs | BIS | ADHD-OCD > OCD<br>BIS total, attentional, and motor.<br>ADHD-OCD had higher mean number of obsessions and compulsions and more frequent symmetry, hoarding, and miscellaneous obsessions and ordering/arranging hoarding compulsions. | Relatively small sample. | BIS total strongly positively associated with comorbid diagnosis of OCD and ADHD. |
| Miyauchi, M., Matsuura, N., Mukai, K., Hashimoto, T., Ogino, S., Yamanishi, K., ... & Matsunaga, H. (2023). A prospective investigation of impacts of comorbid | Japan | 93 OCD (15 ADHD, 25 'ADHD trait', 53 non-ADHD) | MH facility | Clinical | Int – Medication & CBT | BIS | ADHD-OCD > OCD<br>BIS total and all subscales.<br>ADHD-OCD group had an earlier age of onset of OCD; higher | No BIS assessment post-treatment. Excluded ADHD 'trait' |  |
| attention deficit hyperactivity disorder (ADHD) on clinical features and long-term treatment response in adult patients with obsessive-compulsive disorder (OCD). *Comprehensive Psychiatry*, 125, 152401. |  |  |  |  |  |  | levels of depressive, anxious, hoarding, and impulsive symptoms; lower quality of life higher rates of substance/behavioural addiction. Y-BOCS mean improvement rate after 6 months of treatment was significantly lower in the ADHD-OCD group. | group that didn't meet diagnosis for ADHD, but skews data of possibly more impulsive OCD. |  |
| Nagy, N. E., El-Serafi, D. M., Elrassas, H. H., Abdeen, M. S., & Mohamed, D. A. (2020). Impulsivity, hostility and suicidality in patients diagnosed with obsessive compulsive disorder. *International journal of psychiatry in clinical practice*, 24(3), 284-292. | Egypt | 81 OCD (29 SA, 22 SI, 30 no SA/SI) | MH facility | Clinical | Obs | BIS | SA > SI and no SA/SI: BIS attention. SI and no SA/SI > SA: BIS non-planning. | Convenience sampling from one institute. Only SA that resulted in hospitalisation included. Can't assess individuals who completed suicide. Medication not described. | BIS showed no correlation with suicide ideation scale. |
| Namlı, Z., Tamam, L., Demirkol, M. E., Karaytuğ, M. O., & Sun, T. (2023). The relationship among autistic traits, impulsivity, and functionality in patients with obsessive-compulsive disorder. *The Journal of Nervous and Mental Disease*, 211(3), 195-202. | Turkey | 88 OCD<br>90 HC | MH facility, HC were staff relatives | Clinical | Obs | BIS | OCD > HC BIS total, attentional, non-planning. Impulsivity had a partial mediating role in the relationship between autistic traits and disability, and autistic traits had a partial mediating role in the relationship between impulsivity and disability. | Questionnaires used rather than semi-structured interviews - could lead to confusion between autistic/OCD traits. Lack of generalisability given comorbidities excluded. | Significant but weak positive relationship in OCD group between BIS total and WHODAS disability scores, as well as autism quotient scores. |
| Ozdemiroglu, F., Sevincok, L., Sen, G., Mersin, S., Kocabas, O., Karakus, K., & Vahapoglu, F. (2015). Comorbid obsessive-compulsive disorder with bipolar disorder: A distinct form?. *Psychiatry research*, 230(3), 800-805. | Turkey | 61 OCD<br>48 BD<br>32 OCD + BD | MH facility | Clinical | Obs | BIS | BD-OCD > OCD BIS total, motor, and non-planning. Suggests comorbidity of BD and OCD is associated with impulsivity. | Comorbidities excluded. Didn't include BD in active manic episode. |  |
| Parkes, L., Tiego, J., Aquino, K., Braganza, L., Chamberlain, S. R., Fontenelle, L. F., ... & Yücel, M. (2019). Transdiagnostic variations in impulsivity and compulsivity in obsessive-compulsive disorder and gambling disorder correlate with effective connectivity in cortical-striatal-thalamic-cortical circuits. *Neuroimage*, 202, 116070. | USA/<br>Australia | 487 online<br>39 HC<br>34 OCD<br>23 PG | MH facility (OCD), community sample | Clinical/NI | Obs | UPPS-P | GD > OCD and HC impulsive phenotype. In particular, both OCD and GD showed higher scores on the disinhibition phenotype compared to HC. Individuals with high impulsivity showed no differences in effective connectivity, but rather lower inhibitory activity in the self-connection for the medial OFC (meOFC) in the ventral circuit. |  |  |
| Piacentino, D., Pasquini, M., Tarsitani, L., Berardelli, I., Roselli, V., Maraone, A., & Biondi, M. (2016). The association of anger with symptom subtypes in severe obsessive-compulsive disorder outpatients. *Psychopathology*, 49(1), 40-46. | Italy | 57 OCD | MH facility | Clinical | Obs | SVARAD Impulsiveness measure | Found significant correlations between impulsivity dimension and aggressive and sexual obsessions. | Small n, only a sub-scale measure used to report impulsivity. Not homogenous drug treatment, drug treatment not assessed. No comparative group | Impulsivity positively correlated with aggressive and sexual obsessions. |
| Prochazkova, L., Parkes, L., Dawson, A., Youssef, G., Ferreira, G. M., Lorenzetti, V., ... & Yücel, M. (2018). Unpacking the role of self-reported compulsivity and impulsivity in obsessive-compulsive disorder. *CNS spectrums*, 23(1), 51-58. | Australia/<br>Brazil | 103 OCD<br>79 'OCD-likely'<br>35 HC | MH facility, media advertisements, M-Turk survey | Clinical/Survey | Obs | UPPS-P | Clustering done to group OCD depending on their relative compulsivity and impulsivity: low/low, low/high, high/low, high/high. High/high group yielded highest OCD-R scores; low/low had lowest scores. Subgroups held up when adding HC. High/high cluster was exclusively OCD participants. Low/low group comprised mostly of HC. High/low groups showed roughly equal clinical severity, but with opposing primary symptoms. The largest cluster of individuals with OCD was characterised by high impulsivity and low compulsivity. | OCD-likely group sourced online through paying platform – introduces possibility of survey fraud, participant disinterest, biases associated with online participants. |  |
| Sahmelikoglu Onur, O., Tabo, A., Aydin, E., Tuna, O., Maner, A. F., Yildirim, E. A., & Çarpar, E. (2016). Relationship between impulsivity and obsession types in obsessive-compulsive disorder. *International journal of psychiatry in clinical practice*, 20(4), 218-223. | Turkey | 146 OCD<br>80 HC | MH facility | Clinical | Obs | BIS | OCD > HC BIS attention. OCD with sexual, aggressive, religious obsessions > OCD without those characteristics on BIS total and attention. |  |  |
| Schröder, J., Werkle, N., Cludius, B., Jelinek, L., Moritz, S., & Westermann, S. | Germany | 128 OCD<br>(49 TAU,<br>33 iCBT | OCD self-help facility, | Clinical | Int - iCBT | I-8 | Intervention reduced OCD symptoms (compulsions more | Sample well-educated and high rate of | Correlations of Impulsivity with |
| (2020). Unguided Internet-based cognitive-behavioral therapy for obsessive-compulsive disorder: A randomized controlled trial. *Depression and anxiety*, 37(12), 1208-1220. |  | after dropout) | community forums |  |  |  | than obsessions), but no difference in impulsivity. | employment – may limit generalisability . No diagnostic interviews, only self-report. | severity of OCD not reported. |
| Shan, H. D., Liu, Y. F., Zhao, Q., Wang, Y., Wang, Y. M., Cheung, E. F., ... & Wang, Z. (2022). Distinct clinical manifestations of obsessive-compulsive disorder are associated with cortical thickness alteration. *Australian & New Zealand Journal of Psychiatry*, 56(2), 186-196. | China | 89 OCD<br>42 HC | MH facility | Clinical/NI | Obs | BIS | OCD > HC BIS attention and non-planning. Neuroimaging: findings did not survive corrections for multiple comparisons. | Total BIS not reported. Uncorrected neuroimaging results reported in discussion. |  |
| Sharma, P., Gale, T. M., & Fineberg, N. A. (2012). Clinical correlates of tobacco smoking in OCD: A UK, case-controlled, exploratory analysis. *Journal of behavioral addictions*, 1(4), 180-185. | UK | 183 OCD<br>(10 smokers) | MH facility | Clinical | Obs | BIS | OCD smokers > OCD non-smokers BIS total. | Small sample: 10 smokers so used 10 non-smokers as subset. Age and BIS sub-scales not reported. |  |
| Shoval, G., Zalsman, G., Sher, L., Apter, A., & Weizman, A. (2006). Clinical characteristics of inpatient adolescents with severe obsessive-compulsive disorder. *Depression and Anxiety*, 23(2), 62-70. | Israel | 40 OCD<br>118 PsyD<br>53 AD<br>81 CD<br>50 ED<br>87 HC | MH facility | Clinical | Obs | CSPS – Impulsivity measure | HC > OCD impulsivity. Stated that OCD differed in impulsive traits from other diagnostic groups but do not elaborate or indicate any significant differences between groups. | Impulsivity measured by subset of another scale. May be severe OCD sample (inpatients). |  |
| Sohn, S. Y., Kang, J. I., Namkoong, K., & Kim, S. J. (2014). Multidimensional measures of impulsivity in | Korea | 80 OCD<br>76 HC | Not stated | Clinical/NC | Obs | BIS | OCD > HC BIS total, attentional, motor. Increased action and choice impulsivity, |  | BIS scores in OCD and HC did not correlate |
| obsessive-compulsive disorder: cannot wait and stop. *PLoS One*, 9(11), e111739. |  |  |  |  |  |  | and decreased risk-taking propensities were found in OCD. |  | significantly with risk-taking, choice impulsivity, or action impulsivity scores. |
| Stein, D. J., Hollander, E., Simeon, D., & Cohen, L. (1994). Impulsivity scores in patients with obsessive-compulsive disorder. *The Journal of nervous and mental disease*, 182(4), 240-241. | USA | 431 OCD<br>151 HC | MH facility,<br>student controls | Clinical | Obs | BIS | OCD = HC BIS. | HC group students. Not reported whether the 'younger' student group referred to was significantly younger than OCD group - NB in light of finding that BIS increased in younger age. | BIS total, cognitive, and non-planning weakly significantly positively correlated with YBOCS compulsions and YBOCS total. BIS total weakly significantly positively correlated with total aggressive and sexual symptoms. Significant negative correlations of BIS total and target cleaning/checking symptoms and age. BIS not correlated with treatment outcome. |
| Stein, D. J., Mullen, L., Islam, M. N., Cohen, L., de Caria, C. M., & Hollander, E. (1995). Compulsive and impulsive symptomatology in trichotillomania. *Psychopathology*, 28(4), 208-213. |  | 17 OCD<br>12 TTM<br>14 BPD | MH facility | Clinical | Obs | Eysenck 1-7, TPQ | BPD and TTM > OCD impulsiveness. BPD had higher impulsive results than OCD on several measures, but no difference in aggression. | Heterogenous patient sample. Medication not reported. Small sample. |  |
| Storch, E. A., Bussing, R., Jacob, M. L., Nadeau, J. M., Crawford, E., Mutch, P. J., ... & Murphy, T. K. (2015). Frequency and correlates of suicidal ideation in pediatric obsessive-compulsive disorder. *Child Psychiatry & Human Development*, 46, 75-83. | USA | 54 OCD youth | MH facility | Clinical | Obs | SNAP-IV Impulsivity subscale | Parent-rated impulsivity not significantly correlated with suicidal ideation. | Parent-rated scale. No correction for type 1 error used. Small sample that excluded youth with active suicidality or a recent attempt. | No correlation parent-rated impulsivity and youth SI. |
| Summerfeldt, L. J., Hood, K., Antony, M. M., Richter, M. A., & Swinson, R. P. (2004). Impulsivity in obsessive-compulsive disorder: comparisons with other anxiety disorders and within tic-related subgroups. *Personality and Individual Differences*, 36(3), 539-553. | Canada | 40 OCD<br>37 PD<br>24 SP<br>49 HC | MH facility, HC advertisements | Clinical | Obs | BIS | OCD = SP, PD > HC: BIS total and attention.<br>OCD with tics > OCD without tics = SP, PD: BIS total and attention. |  | BIS attention significantly correlated with YBOCS total and obsessions. |
| Tan, O., Metin, B., & Metin, S. (2016). Obsessive-compulsive adults with and without childhood ADHD symptoms. *ADHD Attention Deficit and Hyperactivity Disorders*, 8, 131-138. | Turkey | 83 OCD | MH facility | Clinical | Obs | BIS | 40% of OCD not ADHD diagnosed in childhood but had ADHD symptoms in childhood. Childhood ADHD-OCD > non-childhood ADHD<br>OCD: BIS total and motor. | Medication status not reported, retrospective design. | BIS total significantly positively correlated ADHD rating scale. |
| Tan, O., & Taş, C. (2015). Symptom dimensions, smoking and impulsiveness in obsessive-compulsive disorder. *Psychiatria Danubina*, 27(4), 0-405. | Turkey | 167 OCD | MH facility | Clinical | Obs | BIS | Impulsivity did not differ across OCD symptom subtypes: washing, taboo thoughts, and symmetry/counting/repeating/ordering. | Small N. | In washers who smoke, BIS total positively correlated with BAI. In taboo thought smokers, BIS and Y-BOCS were positively correlated. The severity of addiction does not differ in smokers with OCD across symptom dimensions. |
| Tan, O., & Koroglu, A. F. (2020). Smoking prevalence, nicotine dependence, and impulsivity in obsessive-compulsive disorder. *International Journal of Mental Health and Addiction*, 18(1), 125-137. | Turkey | 314 OCD<br>319 HC | MH facility, snowball sampling HC | Clinical | Obs | BIS | OCD > HC BIS total, attention, non-planning. Current smokers OCD > current smokers HC Never smoked OCD > Never smoked HC BIS total, cognitive, non-planning. Former smokers in both groups didn't differ in impulsiveness. OCD current smokers > OCD former smokers BIS attention. HC group impulsivity did not change with smoking status. | Exclusion of comorbid mood disorders - lack of generalisability . | Positive correlation between BIS total and all subscales and nicotine dependence (Fagerström scores). Stronger correlation in OCD group than HC. |
| Tavares, H., & Gentil, V. (2007). Pathological gambling and obsessive-compulsive disorder: towards a spectrum of disorders of volition. *Brazilian Journal of Psychiatry*, 29, 107-117. | Brazil | 40 OCD<br>40 PG<br>40 HC | MH facility | Clinical | Obs | BIS, TPQ | PG > OCD BIS total and novelty-seeking when controlling for anxiety and depression. |  | Novelty seeking sub-factors correlated strongly among each other and with BIS total. |
| Tumkaya, S., Yucens, B., Mart, M., Tezcan, D., & Kashyap, H. (2021). Multifaceted impulsivity in obsessive-compulsive disorder with hoarding symptoms. *Nordic Journal of Psychiatry*, 75(3), 207-213. | Turkey | 136 OCD<br>94 HC | MH facility | Clinical | Obs | BIS | OCD High hoarding > OCD low hoarding > HC BIS attention. OCD high hoarding > HC BIS motor. OCD high hoarding > OCD low hoarding = HC BIS total. | Many different medications and doses. Low strength of correlations and variance explained indicates | Significant correlations between HRS total and BIS total and attention. HAM-A positively correlated with |
|  |  |  |  |  |  |  |  | factors other than impulsivity may be predictive of hoarding. | BIS total, motor, and attention in OCD. When anxiety included as covariate, HRS of all OCD correlated with BIS total, motor, and attention. |
| Vats, P., Das, B., & Khanra, S. (2020). Serum lipids among drug naïve or drug-free patients with obsessive compulsive disorder and their association with impulsivity: A comparative study. *Indian Journal of Psychological Medicine*, 42(3), 281-289. | India | 40 OCD<br>40 HC | MH facility | Clinical | Obs | BIS | No difference in serum lipids between OCD with high or low impulsivity. | Impulsivity was not reported in the HC group. | Serum HDL levels negatively correlated with BIS total and all subscales in OCD. BIS motor positively correlated with serum VLDL levels. |
| Won Kim, S., & Grant, J. E. (2001). Personality dimensions in pathological gambling disorder and obsessive-compulsive disorder. *Psychiatry research*, 104(3), 205-212. | USA | 41 OCD<br>33 PG<br>40 HC | Pharmacological trials, university advertisements | Clinical | Obs | TPQ | PG > OCD TPQ. | Small, relatively young sample (HC university age; no age difference in groups). Exclusion of comorbidities. | No correlation between severity scores and personality subscales. |
| Xu, T., Gu, Q., Zhao, Q., Wang, P., Liu, Q., Fan, Q., ... & Wang, Z. (2021). Impaired cortico-striatal functional connectivity is related to trait impulsivity in unmedicated patients with obsessive-compulsive disorder. *Journal of Affective Disorders*, 281, 899-907. | China | 36 OCD<br>50 HC | MH facility | Clinical/ NI | Obs | BIS | OCD > HC total, attentional, non-planning. Neuroimaging correlational results did not survive correction for multiple comparisons. | Small sample. | No significant correlations between BIS and YBOCS. |

AD: affective disorder, ADHD: attention deficit hyperactive disorder, AN: anorexia nervosa, AUD: alcohol use disorder, BN: bulimia nervosa, BD: bipolar disorder, BIS: Barratt Impulsivity Scale, BPD: borderline personality disorder, CD: conduct disorder, CSPA: Childhood Suicide Potential Scale, CT: childhood trauma, ED: eating disorder, HC: healthy controls, iCBT: internet-based cognitive behavioural therapy, Int: interventional, IVE: Impulsiveness Venturesomeness Empathy, HRS: Hoarding Rating Scale, LD: learning disability, MH: mental health, NC: neurocognitive, NI: neuroimaging, Obs: observational, PANDAS: Paediatric autoimmune neuropsychiatric disorders associated with streptococcal infections, PD: panic disorder, PG: pathological gambling, PsyD: psychotic disorder, SNAP-IV: Swanson, Nolan, and Pelham – IV Parent Scale, SP: social phobia, SPD: skin-picking disorder, SVARAD: Scale for the Rapid Assessment of Psychopathology, SZD: schizophrenic disorder, TAU: treatment as usual, TPQ: Tridimensional Personality Questionnaire, TS: Tourette syndrome, TTM: trichotillomania,

